# Detailed Quantification of Glomerular Structural Lesions Associates with Clinical Outcomes and Transcriptomic Profiles in Nephrotic Syndrome

**DOI:** 10.1101/2021.09.16.21263706

**Authors:** Jeffrey B. Hodgin, Laura H. Mariani, Jarcy Zee, Q Liu, Abigail R. Smith, Sean Eddy, John Hartman, Habib Hamidi, Joseph P. Gaut, Matthew B. Palmer, Cynthia C. Nast, Anthony Chang, Stephen Hewitt, Brenda W. Gillespie, Matthias Kretzler, Lawrence B. Holzman, Laura Barisoni, for the Nephrotic Syndrome Study Network (NEPTUNE)

## Abstract

The current classification system for focal segmental glomerulosclerosis (FSGS) and minimal change disease (MCD) does not fully capture the complex structural changes in kidney biopsies, nor the clinical and molecular heterogeneity of these diseases. The Nephrotic Syndrome Study Network (NEPTUNE) Digital Pathology Scoring System (NDPSS) was applied to 221 NEPTUNE FSGS/MCD digital kidney biopsies for glomerular scoring using 37 descriptors. The descriptor-based glomerular profiles were used to cluster patients with similar morphologic characteristics. Glomerular descriptors and patient clusters were assessed for association with time to proteinuria remission and disease progression by using adjusted Cox models, and eGFR measures over time by using linear mixed models. Messenger RNA from glomerular tissue was used to assess differentially expressed genes (DEG) between clusters and identify genes associated with individual descriptors driving cluster membership. Three clusters were identified: X (N=56), Y (N=68), and Z (N=97). Clusters Y and Z had higher probabilities of proteinuria remission (HR [95% CI]= 1.95 [0.99, 3.85] and 3.29 [1.52, 7.13], respectively), lower hazards of disease progression 0.22 [0.08, 0.57] and 0.11 [0.03, 0.45], respectively), and greater loss of eGFR over time compared with X. Cluster X had 1920 DEGs compared to Y+Z, which reflected activation of pathways of immune response and inflammation. Six individual descriptors driving the clusters individually correlated with clinical outcomes and gene expression. The NDPSS allows for characterization of FSGS/MCD patients into clinically and biologically relevant categories and uncovers histologic parameters associated with clinical outcomes and molecular signatures not included in current classification systems.

**TRANSLATIONAL STATEMENT:** FSGS and MCD are heterogeneous diseases that manifest with a variety of structural changes often not captured by conventional classification systems. This study shows that a detailed morphologic analysis and quantification of these changes allows for better representation of the structural abnormalities within each patient and for grouping patients with similar morphologic profiles into categories that are clinically and biologically relevant.

## INTRODUCTION

The current histopathology-based classification system of focal segmental glomerulosclerosis (FSGS) and minimal change disease (MCD) does not fully capture the morphologic, molecular, or clinical heterogeneity of these diseases. This classification approach is based on a few, selected parameters (e.g., segmental sclerosis) to derive a diagnosis from the entire biopsy specimen. These diagnostic categories do not capture complex combinations of discrete structural changes and are not derived from the molecular basis of the disease. As a result, the current classification system has limited utility in predicting disease progression, response to therapy, or guiding discovery of new therapeutic approaches. New methodologies for morphologic evaluation of renal biopsies are necessary to extract useful information from the kidney structural changes that can be integrated with clinical and biomarker data to identify participant-specific therapeutic targets and ultimately improve patient care.^1, 2^

The Nephrotic Syndrome Study Network (NEPTUNE) Digital Pathology Scoring System (NDPSS) was developed to capture the morphologic complexity of glomerular diseases.^3^ The NDPSS applies quantitative and semi-quantitative metrics to evaluate the presence and extent of discrete structural changes (“descriptors”) in the glomeruli, tubulointerstitium, and vessels. Morphologic data are objectively collected, quantified and agnostic to conventional categories.^3, 4^ Because of its granularity, this approach is well-suited for integration of this information with other data using systems biology approaches.

To test the hypothesis that quantification of the discrete glomerular structural changes allows for uncovering clinically and biologically relevant information not available using conventional approaches, we applied the NDPSS to kidney biopsies from patients conventionally classified as MCD or FSGS. The resulting patient specific glomerular descriptor-based profiles were used to identify subgroups of participants with morphologic similarities and test associations of these subgroups and individual descriptors with clinical outcomes and molecular phenotype.

## METHODS

### Study Sample

NEPTUNE is a prospective cohort of children and adults enrolled at the time of first clinically indicated biopsy with proteinuria >0.5 mg/mg in phase 1 and >1.5mg/mg in phase 2.^4^ NEPTUNE participants with a digital kidney biopsy in the Digital Pathology Repository (DPR) and a diagnosis of MCD and FSGS were included in this study. FSGS included patients with at least one glomerular lesion of segmental sclerosis, including not otherwise specified, tip, perihilar, cellular and collapsing types.^5^ MCD was further classified into two subsets: MCD (defined as ≥75% effacement with no global sclerosis or global sclerosis expected for age) or MCD-Like (defined as <75% effacement with or without presence of global sclerosis exceeding that expected for age or ≥75% effacement with global sclerosis exceeding that expected for age).^6^ According to the NEPTUNE protocol, kidney biopsies are not further characterized based on etiology (i.e., primary, secondary and adaptive). Clinical demographics, medical history, kidney failure status, medication exposures and local laboratory data, were collected at each study visit. Additional details can be found in S1 Supplementary Methods.

### NEPTUNE Digital Pathology Scoring System (NDPSS) and Patient Clustering by Morphologic Profiles

Glomeruli were annotated (enumerated) across all levels (Figure 1 A-B) and each glomerulus was scored by study pathologists using all levels, according to the NDPSS, so that each glomerular profile was the result of the sum of all individual descriptors observed^3, 7^ (Figure 1C). A descriptor reference manual and an electronic scoring sheet was used to record the scoring of 37 glomerular descriptors through all biopsy sections and stains (S2 Supplementary Table 1).^3, 8^ These descriptors represent a comprehensive list of structural and cellular abnormalities that can be seen in a glomerulus. The overall presence of each descriptor in a participant’s biopsy was quantified by calculating the percentage of a participant’s glomeruli with each descriptor present. We applied Ward’s hierarchical clustering algorithm to cluster participants into subgroups (detailed in S1). The final cluster solution was compared with conventional morphologic classifications (MCD/MCD-Like and FSGS subtypes) ^5, 9^ using a Sankey diagram. To identify the strongest individual descriptors driving cluster membership, we used elastic net penalized multinomial regression with a penalization mixing parameter of α=0.01. We also assessed whether a subset of descriptors could predict cluster membership by repeating the model with only the top 5, 10, 15, 20, 25 and 30 descriptors and comparing prediction accuracy across models.

**Table 1.**
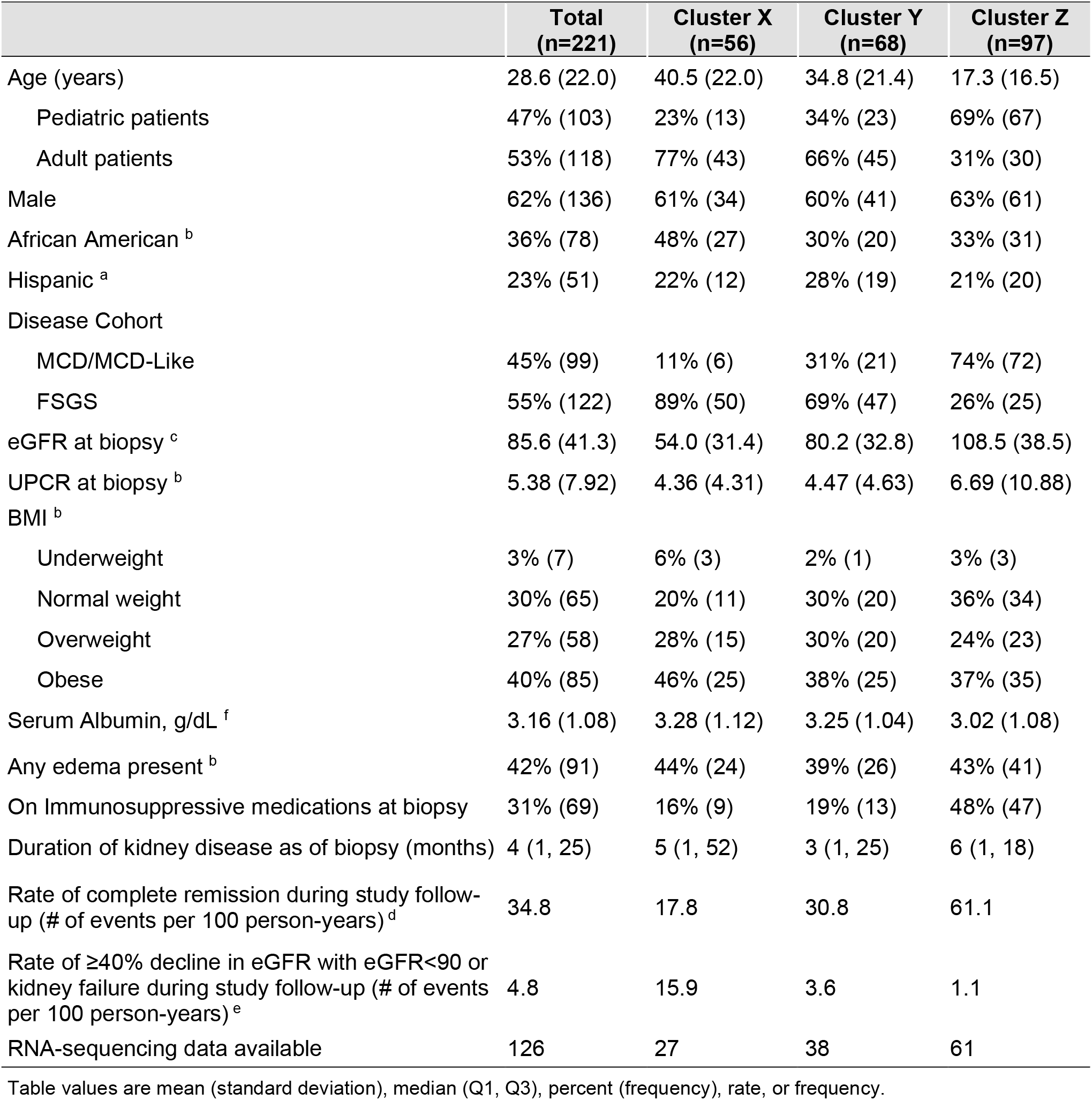
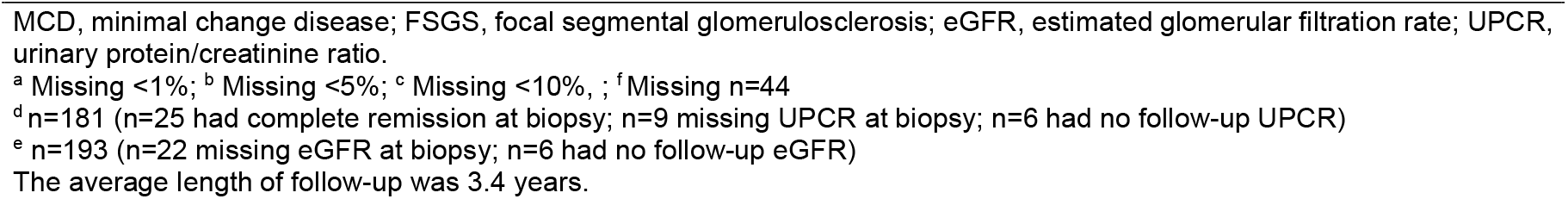
Patient characteristics overall and by glomerular cluster.

**Figure 1:**
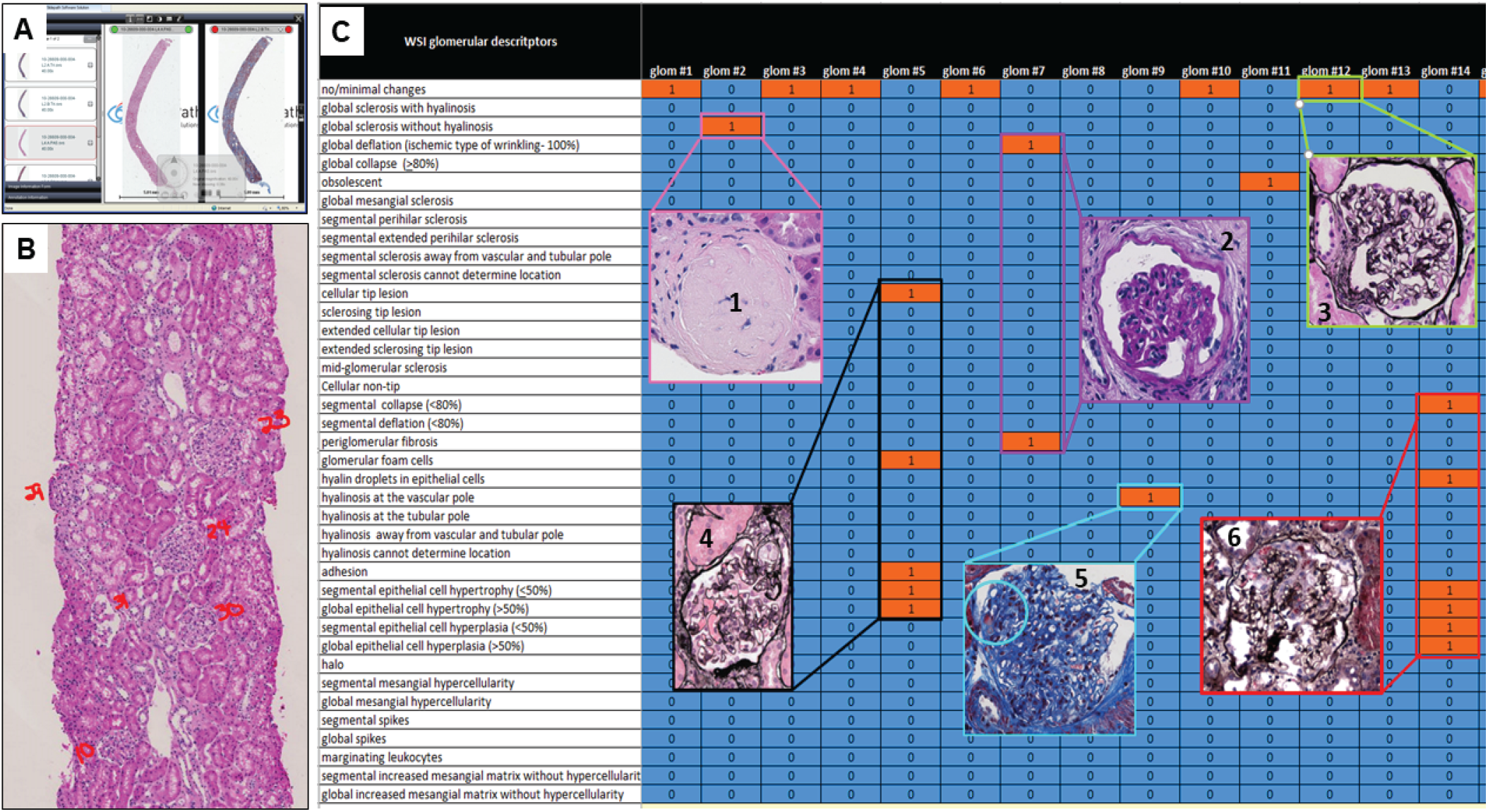
The NEPTUNE Digital Pathology Protocol (NDPP) and Scoring System (NDPSS). Shown are examples of visualization of whole slide images (A), annotation (B) and scoring (C). According to the NDPP, renal biopsies are scanned into whole slide images (A) and manually enumerated (B). Scoring pathologists record all descriptors observed at each level for each of the enumerated glomeruli using an electronic scoring sheet (C). Shown from left to right and top to bottom, are examples of six glomeruli characterized by different descriptors: 1. descriptor global sclerosis without hyalinosis (hematoxylin & eosin); 2. descriptor global deflation and periglomerular fibrosis were selected to profile this glomerulus (periodic acid Schiff); 3. when a glomerulus was histologically unremarkable, the descriptor no/minimal changes was selected (silver stain); 4. descriptors cellular tip lesion, glomerular foam cell, segmental visceral epithelial cell hypertrophy and segmental hyperplasia (silver stain). While in this level the glomerulus shows only segmental visceral epithelial cell hypertrophy, if in a different level reveals global hypertrophy was noted, then both segmental and global visceral epithelial cell hypertrophy are recorded; 5. descriptor hyalinosis at the vascular pole is selected in this glomerulus (trichrome stain), note the absence of accumulation of extracellular matrix (sclerosis); 6. descriptors segmental collapse, segmental visceral epithelial cell hypertrophy, segmental visceral epithelial cell hyperplasia and hyaline droplets (silver stain).

### Clinical Outcomes Analysis

NEPTUNE participants are followed prospectively with study visits three times per year in the first year and twice yearly thereafter, up to 5 years. Clinical outcomes of interest included **(1)** time from biopsy to complete remission of proteinuria, defined as UPCR <0.3 mg/mg, **(2)** time from biopsy to a composite disease progression outcome of at least 40% decline in eGFR ^10^ with eGFR <90 mL/min/1.73m^2^ or kidney failure, and **(3)** longitudinal eGFR over time. For the two time-to-event outcomes, we used Cox proportional hazards models, and for longitudinal eGFR, we used a linear mixed model with random intercept and random slope for each participant to account for repeated measures. Interactions between time from biopsy and cluster groups were tested in the longitudinal eGFR model to assess whether eGFR slope differed across clusters.

We performed both unadjusted and sequentially adjusted models between cluster groups and outcomes. For adjusted models, we first adjusted for demographics and clinical characteristics. If outcomes had sufficient numbers of events or observations to avoid overfitting, we adjusted for age, sex, self-reported black race, Hispanic ethnicity, eGFR at biopsy, UPCR at biopsy, kidney disease duration as of biopsy, and any immunosuppression use at or within 30 days prior to biopsy; if there were not enough events or observations to avoid overfitting, we used backward selection to choose adjustment covariates. Then, we additionally adjusted for participant disease diagnosis (MCD/MCD-Like vs. FSGS). Finally, we used unadjusted models to assess associations between each individual glomerular descriptor and each outcome. P-values were controlled for false discovery rate (FDR) using the Benjamini and Hochberg linear step-up method.

### Molecular Data and Analysis

Messenger RNA (mRNA) expression from RNA sequencing of microdissected glomeruli was available for 126 of the MCD/MCD-Like and FSGS participants (N=56 and N=70, respectively). (detailed in S1). Glomerular gene expression from cluster X was compared to the clusters Y+Z using Significance Analysis of Microarray (SAM). Differentially expressed genes (DEGs) were defined as having fold change of greater than 1.3 and false discovery rate (FDR)-corrected q-value less than 5%. In addition, Pearson correlation coefficient was calculated between each glomerular RNA-seq-derived gene expression value and log10-transformed descriptor percentage score (after descriptors with 0% were transformed to 1% to avoid undefined values), with adjustment for multiple testing using q-value less than 5% as the criterion for statistical significance. DEGs for the cluster comparison and the transcripts that correlated with individual descriptors, were analyzed for enrichment of canonical pathways and upstream regulators using the Ingenuity Pathway Analysis Software Suite.^11^ An activation z-score was derived from the direction of gene regulation observed to the expected direction of gene regulation to infer the activation state of a putative upstream regulator,^11^ with adjustment for multiple testing using Bonferroni less than 5% as the criterion for statistical significance. Upstream regulators were identified using both enrichment as well as direction of expression change with known cause and effect relationships. In addition, we generated a list of genes that are specific to known renal and non-renal cell types using single cell RNA-seq profiles derived from three tumor nephrectomy datasets, a subset of data from Menon et al.^12^. The expression values of the subset of cell-specific genes were extracted from the glomerular compartment bulk transcriptome data and were labeled according to cell type.^13^ A heatmap of cell type specific transcripts was generated based on the VOOM transformed log expression in which participants were ordered based on the pathology cluster assignment and transcripts were grouped by cell type.

## RESULTS

### Cluster Analysis Results

Three clusters, X, Y and Z, were found using participants’ glomerular descriptors in the hierarchical clustering algorithm (Figure 2A). Cluster X (56 participants) had the most frequent representation of structural changes, followed by Cluster Y (68 participants), and Cluster Z (97 participants) (Figure 2B). Cluster X had the highest prevalence of glomeruli with global and segmental glomerulosclerosis, periglomerular fibrosis, adhesions, and podocyte hypertrophy. Cluster Y had a moderate percentage of globally and segmentally sclerotic glomeruli, adhesion, and podocyte hypertrophy. In contrast, Cluster Z had many glomeruli with no or minimal changes and low prevalence of other descriptors. When we conducted a sensitivity analysis using unscaled data the cluster results remained unchanged.

**Figure 2:**
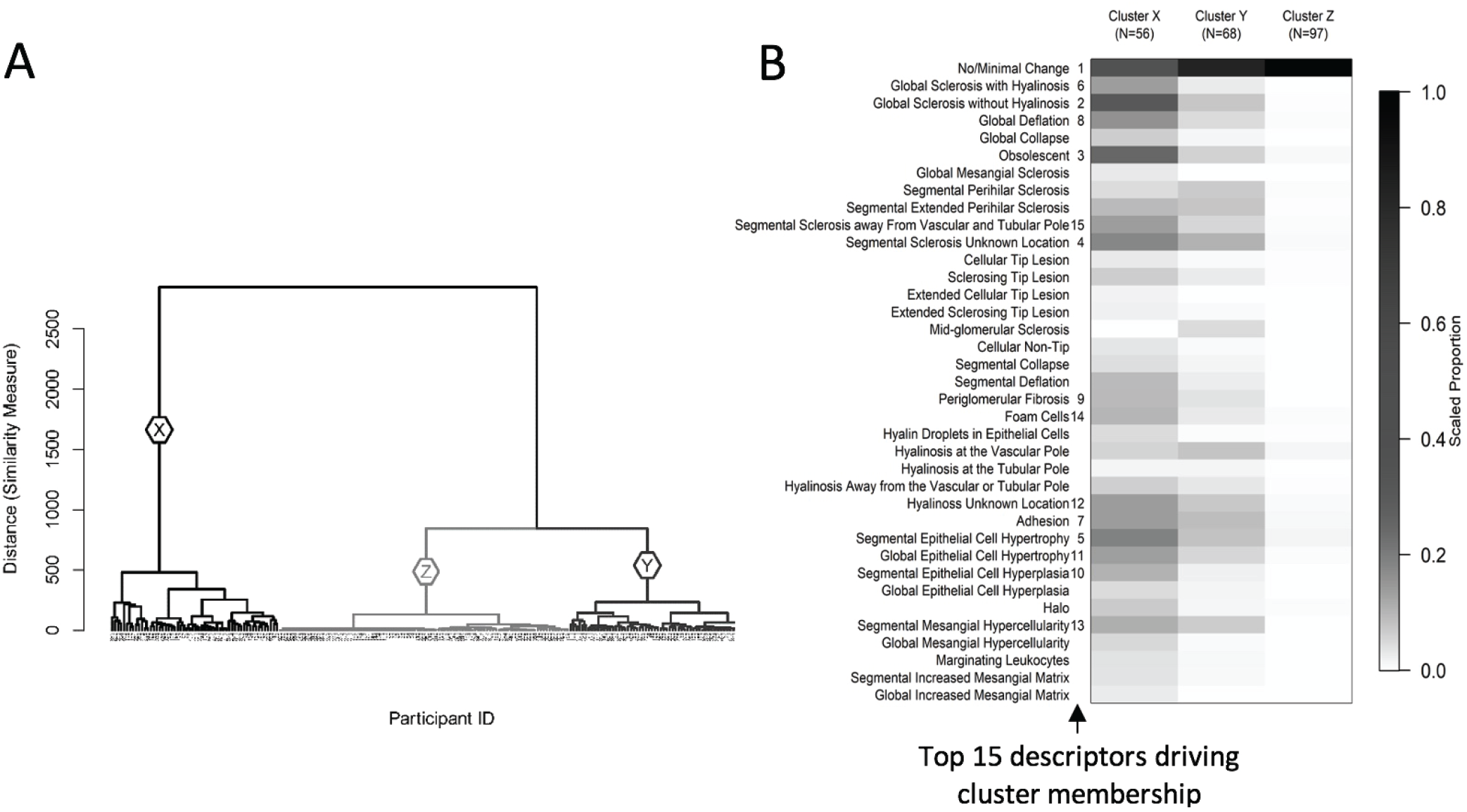
Hierarchical clustering of 221 NEPTUNE participants with FSGS and MCD based on 39 glomerular descriptors. The dendrogram (A) shows 3 unique clusters X (N=56), Y (N=68), and Z (N=97). The heat map (B) shows the mean value of each descriptor by cluster after scaling each descriptor to 0-1 based on the observed range (i.e. the descriptor’s observed minimum is scaled to 0 and maximum to 1). The darker cells therefore represent greater percentages of glomeruli with each descriptor relative to others in the sample. The numbers 1-15 to the right side of descriptor labels indicate variable importance in driving cluster membership, obtained from variable entry order in a penalized multinomial regression model.

The accuracy of cluster membership using fewer descriptors was assessed based on a hierarchy of the top descriptors identified using penalized multinomial regression. Rankings of the top descriptors driving cluster membership are shown in Figure 2B. Using only the top 5 descriptors yielded 81.4% accuracy, the top 10 gave 83.3% accuracy, the top 15 had 89.1% accuracy, the top 20 had 88.2% accuracy, the top 25 had 89.6% accuracy, and the top 30 had 88.7% accuracy. Compared with the prediction accuracy of 88.2% using all 37 glomerular descriptors, the prediction accuracy began to level off with 15 descriptors.

Participants in Clusters X and Y had higher proportions of adults (77% and 66%, respectively) and FSGS participants (89% and 69%, respectively), whereas Cluster Z participants had more pediatric participants (69%) and MCD/MCD-Like participants (74%) (Table 1). Cluster X had the highest percentage of African American participants (48%), compared to Clusters Y and Z (30% and 33%, respectively). Cluster Z participants had the highest mean eGFR and UPCR and had a greater proportion on immunosuppressive medications at biopsy. When compared to conventional diagnostic classification, each descriptor-derived cluster contained participants with MCD/MCD-Like and FSGS subtypes (Figure 3). While over half of Cluster Z patients had MCD/MCD-Like, there was little other concordance between cluster groups and conventional disease classifications.

**Figure 3:**
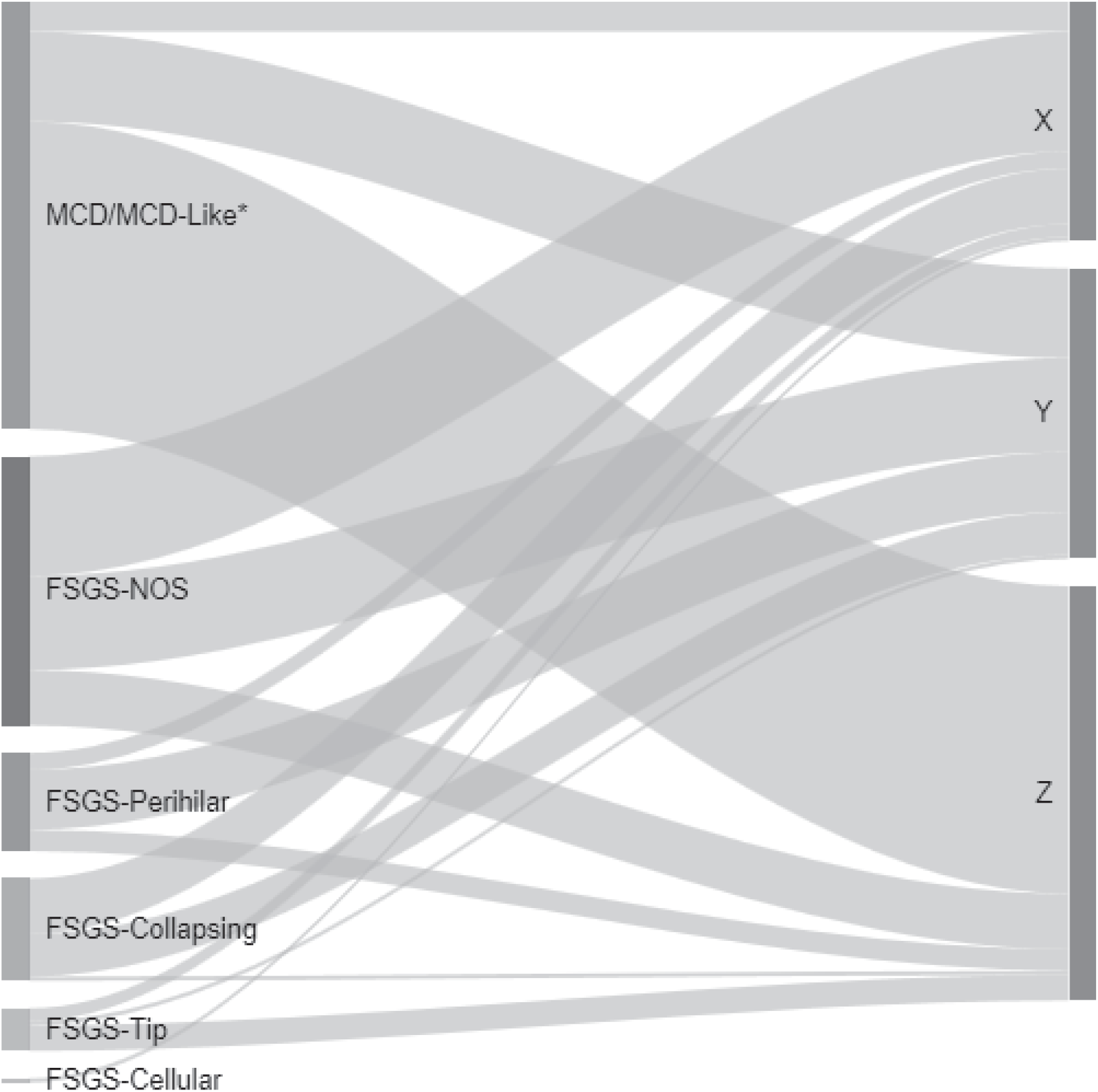
Sankey diagram showing relationships between conventional classification categories (left) and cluster membership (right). The width of each ribbon-shaped band is proportional to the number of patients in that band. *MCD refers to cases with ≥75% effacement with no global sclerosis or global sclerosis expected for age, and MCD-Like refers to cases with <75% effacement with or without presence of global sclerosis exceeding that expected for age or ≥75% effacement with global sclerosis exceeding that expected for age.

#### Clinical outcomes

Cluster X demonstrated both the lowest probability of proteinuria remission and highest probability of disease progression (Figure 4). While Clusters Y and Z had similarly low probabilities of disease progression, Cluster Z had higher probabilities of remission of proteinuria. Cox models gave similar results, both unadjusted and after adjusting for baseline demographics and clinical characteristics (Table 2). In adjusted models, Clusters Y and Z had higher probabilities of proteinuria remission compared to Cluster X (HR [95% CI] = 1.95 [0.99, 3.85] and 3.29 [1.52, 7.13], respectively) and lower probabilities of disease progression (HR [95% CI] = 0.22 [0.08, 0.57] and 0.11 [0.03, 0.45], respectively).

**Table 2:**
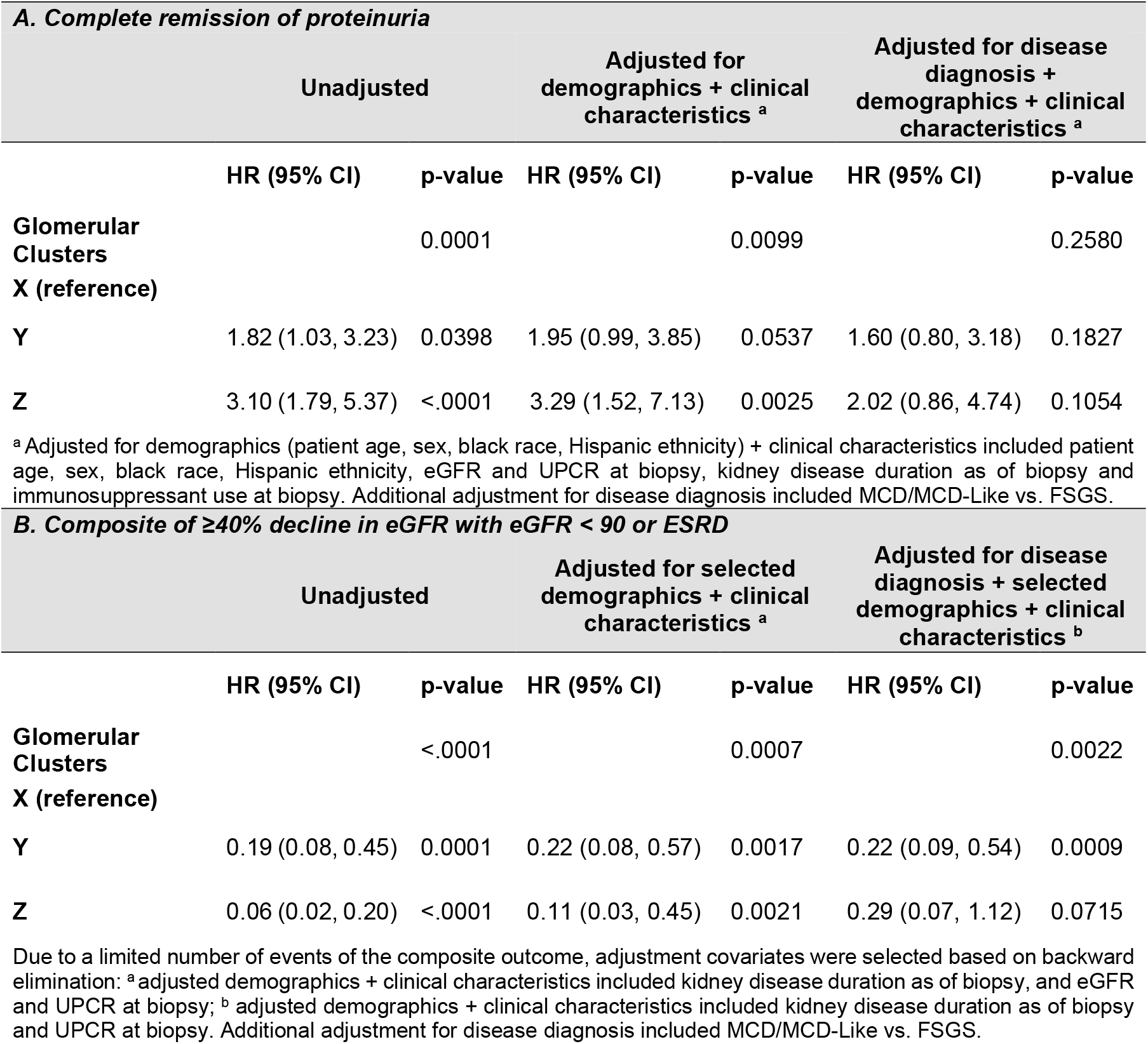
Associations between glomerular clusters and time-to-event outcomes from Cox models, unadjusted and adjusted for demographics, clinical characteristics, and disease diagnosis.

**Figure 4:**
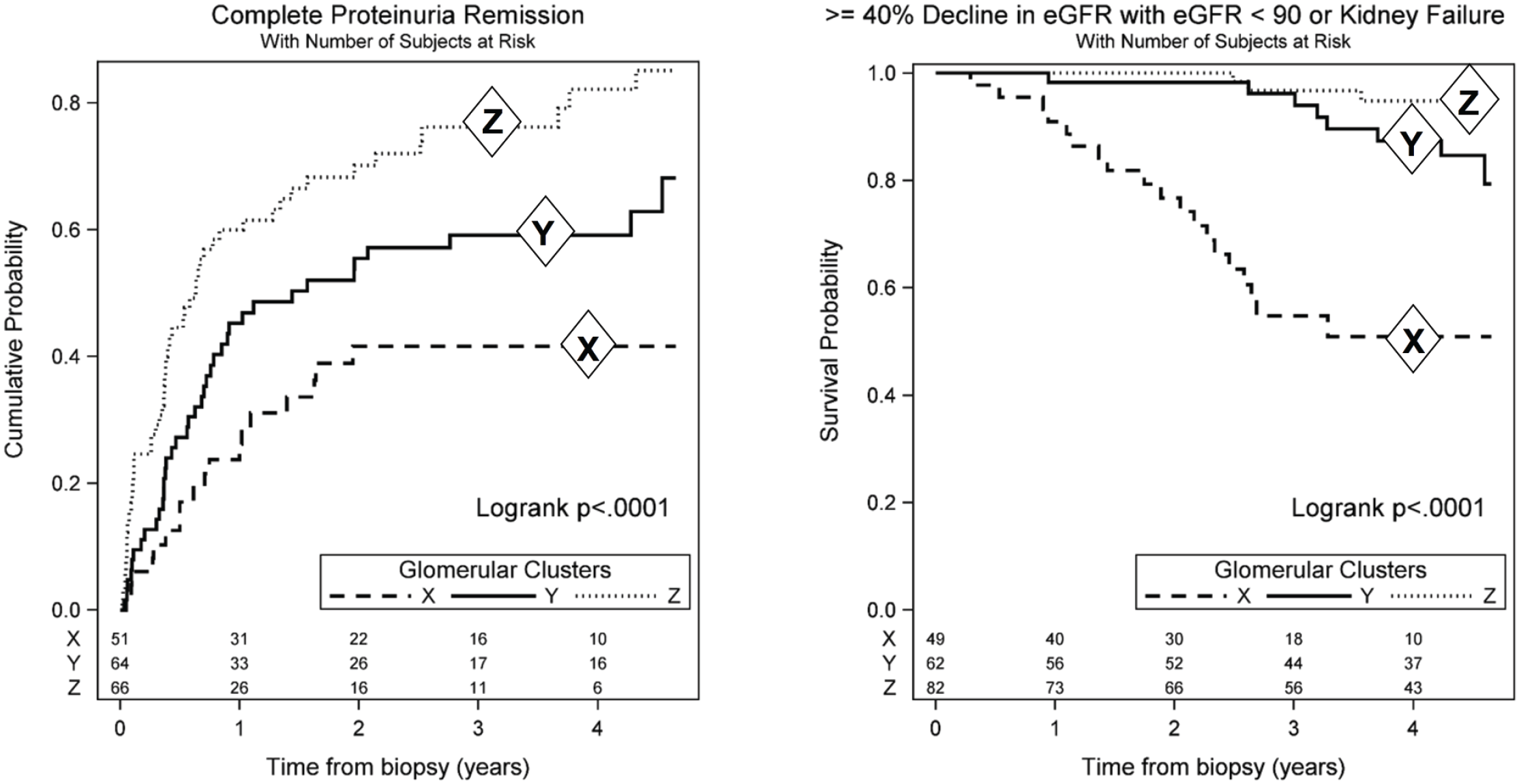
Kaplan-Meier curves for each of the three clusters showing cumulative probability of complete proteinuria remission (left) and the survival probability of ≥40% decline in eGFR with eGFR <90 or kidney failure (right) by clusters. Cluster X had the lowest (worst) probability of proteinuria remission and highest (worst) probability of disease progression, while Cluster Z had the highest proteinuria remission probabilities and Clusters Y and Z had similar disease progression probabilities.

For the longitudinal eGFR outcome and after adjusting for baseline demographics and clinical characteristics, on average, Clusters Y and Z had 13.2 (95% CI: 3.4 to 23.0) and 18.2 (95% CI: 7.6 to 28.9) higher eGFR at biopsy compared to Cluster X, respectively (Table 3). In addition, Clusters Y and Z had slower eGFR decline rates (changes in eGFR of -2.5 [95% CI -4.2 to -0.7] per year and -1.5 [95% CI -3.2 to 0.1], respectively) as compared to Cluster X (change in eGFR of -5.5 [95% CI -7.9 to -3.2] per year) (Table 3, Figure 5).

**Table 3:** Associations between glomerular clusters and longitudinal eGFR from linear mixed models, unadjusted and adjusted for demographics, clinical characteristics, and disease diagnosis.

|  | Unadjusted |  | Adjusted for demographics + clinical characteristics <sup>a</sup> |  | Adjusted for disease diagnosis + demographics + clinical characteristics <sup>a</sup> |  |
| --- | --- | --- | --- | --- | --- | --- |
|  | Estimate (95% CI) | p-value | Estimate (95% CI) | p-value | Estimate (95% CI) | p-value |
| **Glomerular Clusters** |  | <.0001 |  | 0.0029 |  | 0.0175 |
| **X (reference)** |  |  |  |  |  |  |
| **Y** | 21.4 (10.1, 32.7) | 0.0002 | 13.2 (3.4, 23.0) | 0.0086 | 12.3 (2.5, 22.1) | 0.0137 |
| **Z** | 45.8 (35.2, 56.4) | <.0001 | 18.2 (7.6, 28.9) | 0.0008 | 15.0 (3.9, 26.1) | 0.0083 |
| **eGFR slope (per year) by Cluster** |  | 0.0197 |  | 0.0196 |  | 0.0197 |
| **X** | -5.8 (-8.1, -3.5) | <.0001 | -5.5 (-7.9, -3.2) | <.0001 | -5.5 (-7.8, -3.2) | <.0001 |
| **Y** | -2.3 (-4.0, -0.6) | 0.007 | -2.5 (-4.2, -0.7) | 0.0060 | -2.5 (-4.2, -0.7) | 0.0060 |
| **Z** | -2.0 (-3.5, -0.4) | 0.0129 | -1.5 (-3.2, 0.1) | 0.0679 | -1.5 (-3.1, 0.1) | 0.0723 |

Notes: Significant interactions between clusters and time since biopsy were detected in all models.
<sup>a</sup> Adjusted demographics + clinical characteristics included patient age, sex, black race, Hispanic ethnicity, eGFR and UPCR at biopsy, kidney disease duration as of biopsy, and immunosuppressant use at biopsy. Additional adjustment for disease diagnosis included MCD/MCD-Like vs. FSGS.

**Figure 5:**
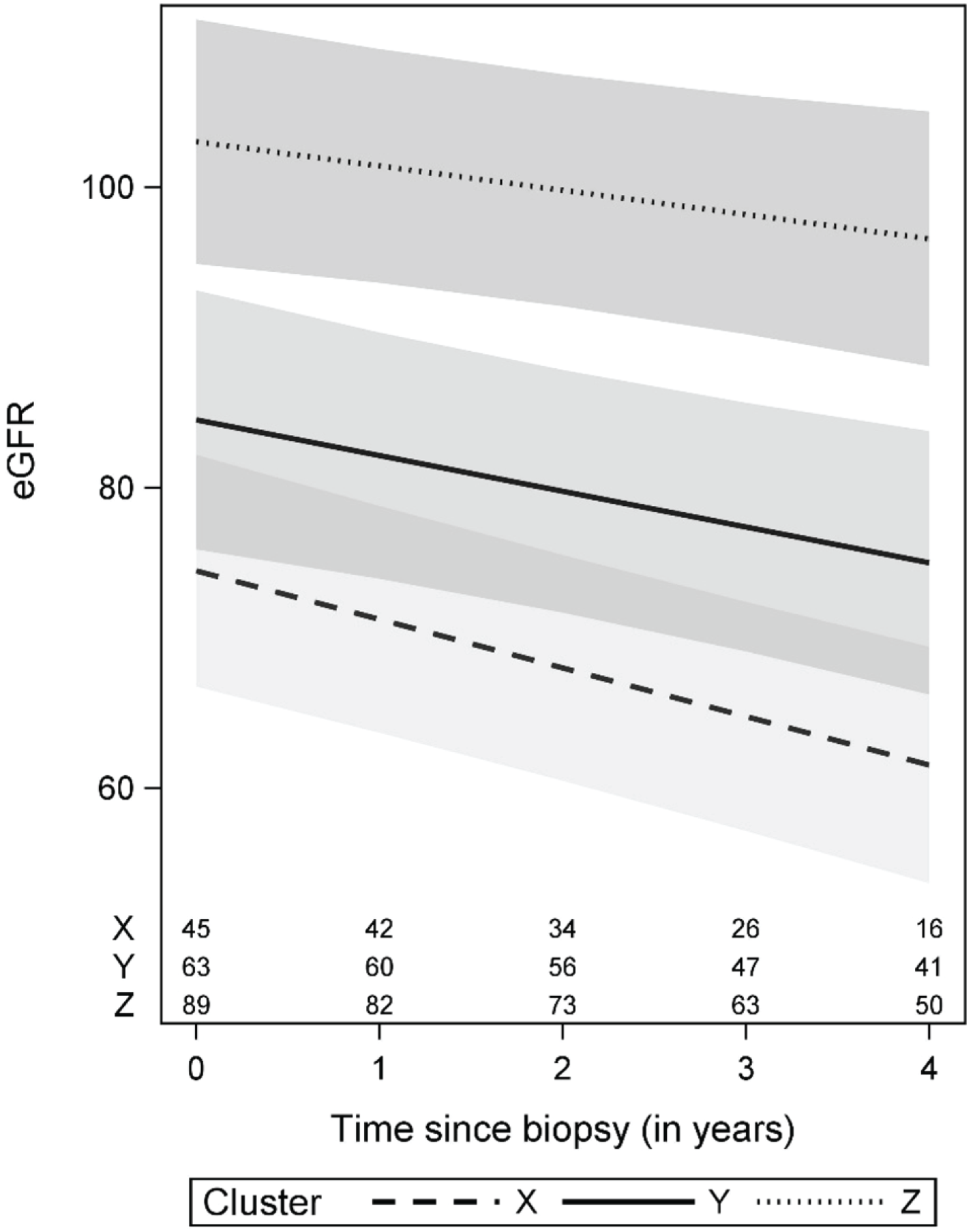
Estimated eGFR trajectories by cluster from longitudinal eGFR models. eGFR trajectories in this figure were estimated for a patient with average characteristics: 28 years old, male, non-black, non-Hispanic, with biopsy UPCR > 3 mg/mg, kidney disease duration > 4 months as of biopsy and not on immunosuppressive medications at biopsy. Cluster-specific averages were used for eGFR at biopsy (57, 79, and 109 mL/min/1.73m2 for Clusters X, Y, and Z, respectively). Confidence bands are based on 95% pointwise confidence intervals. Numbers at the bottom of the graph represent the number of patients available for analysis at each time point.

After additionally adjusting for disease diagnosis, cluster groups were still significantly associated with disease progression (overall p-value=0.002, Table 2) and eGFR change over time (overall p-value for the interaction between cluster groups and time from biopsy=0.0197, Table 3). The magnitude of the association between cluster groups and eGFR change over time was similar after adjustment for disease diagnosis versus before, while effect estimates for cluster groups on time-to-event outcomes were slightly dampened. Unadjusted associations between individual descriptors and outcomes for the top 15 glomerular descriptors that drive cluster membership (Table 4 and S3 Supplementary Table 2) show strong association for most global obliteration descriptors, several types of segmental sclerosis, segmental deflation, hyalinosis, and adhesion.

**Table 4.** Unadjusted associations of the top 15 individual glomerular descriptor and outcomes. Effect estimates presented are for a one-tenth increase in the observed range (observed max – observed min) of a descriptor.

|  |  | % of Glomeruli with Descriptor |  |  |  |  | Complete Remission |  | Composite Progression |  | Longitudinal eGFR |  |
| --- | --- | --- | --- | --- | --- | --- | --- | --- | --- | --- | --- | --- |
|  |  | %N with 0 glomeruli with descriptor | Median (IQR) among non-zero values* | Mean | Min | Max | HR (95% CI) | P-value** | HR (95% CI) | P-value** | Estimate (95% CI) | P-value** |
| No/Minimal Changes | 1 | 0% | 90.9 (68.4 - 100.0) | 78.6 | 6.3 | 100 | 1.25 (1.14, 1.37) | **<.0001** | 0.71 (0.64, 0.80) | **<.0001** | 8.0 (6.7, 9.4) | **<.0001** |
| Global Sclerosis without Hyalinosis | 2 | 59% | 12.2 (6.1 - 23.9) | 6.7 | 0 | 66.7 | 0.82 (0.71, 0.95) | **0.0218** | 1.30 (1.14, 1.48) | **0.0007** | -10.3 (-12.6, -8.0) | **<.0001** |
| Obsolescent | 3 | 62% | **7.7 (3.7 - 21.1)** | 6.2 | 0 | 73.3 | 0.69 (0.56, 0.85) | **0.0027** | 1.40 (1.24, 1.59) | **<.0001** | -9.0 (-11.6, -6.5) | **<.0001** |
| Segmental Sclerosis Unknown Location | 4 | 59% | 9.1 (4.8 - 14.9) | 4.9 | 0 | 61.5 | 0.64 (0.51, 0.80) | **0.0004** | 1.25 (1.09, 1.42) | **0.0042** | -7.3 (-10.3, -4.3) | **<.0001** |
| Segmental Visceral Epithelial Cell Hypertrophy | 5 | 55% | 6.7 (3.9 - 15.8) | 5.6 | 0 | 70.6 | 0.88 (0.75, 1.03) | 0.2007 | 1.17 (1.00, 1.36) | <sup>0.0966</sup><sub>#</sub> | -4.3 (-7.3, -1.4) | **0.0139** |
| Global Sclerosis with Hyalinosis | 6 | 81% | 6.7 (4.0 - 14.3) | 2.1 | 0 | 50 | 0.71 (0.52, 0.97) | <sup>0.0653</sup><sub>#</sub> | 1.37 (1.17, 1.61) | **0.0006** | -10.3 (-13.8, -6.8) | **<.0001** |
| Adhesion | 7 | 61% | 6.7 (4.2 - 10.5) | 3.8 | 0 | 57.9 | 0.69 (0.55, 0.87) | **0.0057** | 1.37 (1.20, 1.55) | **<.0001** | -7.5 (-10.6, -4.4) | **<.0001** |
| Global Deflation | 8 | 76% | 6.0 (3.2 - 14.3) | 2.5 | 0 | 43.8 | 0.77 (0.62, 0.94) | **0.0333** | 1.29 (1.11, 1.50) | **0.0033** | -9.8 (-13.0, -6.6) | **<.0001** |
| Periglomerular Fibrosis | 9 | 75% | 7.7 (3.9 - 14.3) | 2.7 | 0 | 75 | 0.51 (0.34, 0.78) | **0.0059** | 1.22 (1.01, 1.47) | <sup>0.0874</sup><sub>#</sub> | -14.1 (-18.6, -9.6) | **<.0001** |
| Segmental Visceral Epithelial Cell Hyperplasia | 10 | 84% | 7.0 (3.5 - 12.8) | 1.7 | 0 | 52.9 | 0.90 (0.72, 1.14) | 0.4986 | 1.36 (1.06, 1.74) | **0.0372** | -8.3 (-13.1, -3.5) | **0.0032** |
| Global Visceral Epithelial Cell Hypertrophy | 11 | 79% | 6.2 (3.2 - 13.5) | 1.9 | 0 | 35.3 | 0.99 (0.87, 1.13) | 0.9230 | 1.19 (1.00, 1.43) | 0.1097 | -6.3 (-9.8, -2.9) | **0.0019** |
| Hyalinosis Unknown Location | 12 | 77% | 4.7 (3.3 - 9.1) | 1.6 | 0 | 27.3 | 0.66 (0.51, 0.84) | **0.0036** | 1.23 (1.06, 1.44) | **0.0192** | -5.5 (-8.7, -2.3) | **0.0033** |
| Segmental Mesangial Hypercellularity | 13 | 80% | 9.1 (4.8 - 18.2) | 2.9 | 0 | 60 | 1.03 (0.91, 1.16) | 0.7725 | 1.13 (0.94, 1.34) | 0.2936 | 2.0 (-1.2, 5.2) | 0.3202 |
| Foam Cells | 14 | 77% | 6.7 (3.6 - 12.8) | 2.7 | 0 | 75 | 0.92 (0.77, 1.09) | 0.4388 | 1.13 (0.94, 1.35) | 0.2936 | -3.6 (-7.4, 0.2) | 0.1183 |
| Segmental Sclerosis<br>away From Vascular<br>and Tubular Pole | 15 | 87% | 3.6 (2.4 - 6.5) | 0.6 | 0 | 10.5 | 0.83 (0.71, 0.97) | **0.0450** | 1.26 (1.11, 1.44) | **0.0023** | -4.2 (-6.9, -1.5) | **0.0090** |

\*Median (IQR) among non-zero values were based on only a few participants for descriptors with >95% zero values. \*\*P-values were controlled for false discovery rate (FDR) using the Benjamini and Hochberg linear step-up method. # Covariates where 95% CI indicated statistically significant at level 0.05, but FDR adjusted p-values were statistically insignificant.

### Molecular Analysis Results

Glomerular gene expression for participants in the cluster with the worst clinical outcomes (X), was compared to the other two clusters combined to obtain a differentially expressed gene (DEG) list of 1920 DEGs. For insight into the biological and molecular mechanisms, we subjected the DEGs to canonical pathway and upstream regulator enrichment analysis. Table 5 shows the top 25 canonical pathways (pathways predicted to be enriched based on DEGs and ordered by p-value) associated with cluster comparison X vs Y+Z. Table 5 reflects several pathways important for inflammation and immune responses in the kidney, such as dendritic cell maturation, B cell receptor signaling, communication between innate and adaptive immune cells, and role of pattern recognition receptors.^14^ A pathway network of the top 25 canonical pathways is shown in Figure 6 to demonstrate the number of genes shared across pathways, number of enriched genes within each pathway and pathway activation or inhibition. Most of these pathways are predicted to be activated in Cluster X with the exception of ‘LXR/RXR activation’. Table 6 shows the top 25 upstream regulators predicted to be causally upstream of the associated DEGs from X vs Y+Z. The list includes several cytokines also known to be associated with kidney disease progression, such as tumor necrosis factor (TNF), IFN-Gamma, and interleukin-1 beta (IL1B), and growth factors such as transforming growth factor beta (TGFB).^15-17^ Differential regulation of 15 genes chosen from those that predict the activation of these top 4 upstream regulators was validated by the Affymetrix ST 2.1 platform for 77 cases of cluster X relative to Y+Z (14 vs 63 respectively) (S4 Supplementary Table 3 and Supplemental Methods). All 15 DEGs match in direction of fold change of glomerular expression between RNA sequencing and Affymetrix platforms, and differential expression was significant to FDR < 0.05 for 13 genes, with the remaining at FDR of 0.052 and 0.054.

**Table 5.** Top 25 canonical pathways predicted from cluster X vs Y+Z differentially expressed genes.

| **Ingenuity Canonical Pathways** | **Activation**<br>**z-score** | **-log**<br>**(p-value)** |
| --- | --- | --- |
| Phagosome Formation |  | 14.1 |
| Hepatic Fibrosis / Hepatic Stellate Cell Activation |  | 12.7 |
| Dendritic Cell Maturation | 5.47 | 9.78 |
| GP6 Signaling Pathway | 4.70 | 8.46 |
| Granulocyte Adhesion and Diapedesis |  | 8.08 |
| Neuroinflammation Signaling Pathway | 4.90 | 7.38 |
| Role of Macrophages, Fibroblasts and Endothelial Cells in Rheumatoid Arthritis |  | 7.35 |
| B Cell Receptor Signaling | 4.27 | 7.21 |
| Communication between Innate and Adaptive Immune Cells |  | 7.18 |
| Th2 Pathway | 2.13 | 7.17 |
| Role of Pattern Recognition Receptors in Recognition of Bacteria and Viruses | 3.84 | 7.07 |
| Altered T Cell and B Cell Signaling in Rheumatoid Arthritis |  | 7.04 |
| Th1 and Th2 Activation Pathway |  | 6.82 |
| Atherosclerosis Signaling |  | 6.75 |
| Role of NFAT in Regulation of the Immune Response | 4.60 | 6.39 |
| Tec Kinase Signaling | 3.90 | 6.32 |
| Primary Immunodeficiency Signaling |  | 6.19 |
| PI3K Signaling in B Lymphocytes | 4.27 | 6.08 |
| LPS/IL-1 Mediated Inhibition of RXR Function | 2.50 | 6.06 |
| TREM1 Signaling | 4.02 | 5.97 |
| p53 Signaling | 1.88 | 5.79 |
| FXR/RXR Activation |  | 5.75 |
| Acute Myeloid Leukemia Signaling | 0.69 | 5.63 |
| CD28 Signaling in T Helper Cells | 2.83 | 5.35 |
| LXR/RXR Activation | -2.84 | 5.08 |

Top 25 based on p-value. For reference, $-\log(0.05/1000)=4.30$ , so all p-values shown in the table are statistically significant at level 0.05 even after a conservative Bonferroni correction for 1000 tests. The Ingenuity Pathway Analysis database lacked enough data to calculate activation z-scores for some canonical pathways. These are left blank.

**Table 6.** Top 25 upstream regulators predicted from cluster X vs Y+Z differentially expressed genes.

| Upstream Regulator (Gene ID) | Expression Fold Change | Molecule Type | Activation z-score | p-value of overlap |
| --- | --- | --- | --- | --- |
| TNF | 1.518 | cytokine | 10.128 | 6.47E-55 |
| IFNG |  | cytokine | 8.61 | 5.47E-56 |
| IL1B | 1.667 | cytokine | 8.125 | 1.15E-31 |
| TGFB1 | 1.321 | growth factor | 7.435 | 1.71E-59 |
| IL6 |  | cytokine | 6.537 | 9.25E-36 |
| ERBB2 | -1.035 | kinase | 6.053 | 6.39E-37 |
| IL2 |  | cytokine | 6.279 | 3.33E-20 |
| PTGER2 | 1.314 | g-protein coupled receptor | 4.816 | 2.83E-22 |
| SPI1 | 1.664 | transcription regulator | 4.295 | 1.17E-28 |
| FOXO1 | -1.076 | transcription regulator | 4.362 | 2.48E-19 |
| STAT3 | 1.121 | transcription regulator | 3.713 | 8.16E-22 |
| CSF3 |  | cytokine | 2.925 | 1.36E-19 |
| IL4 |  | cytokine | 4.741 | 4.06E-28 |
| IL13 |  | cytokine | 3.143 | 6.04E-20 |
| RARA | 1.058 | nuclear receptor |  | 2.35E-21 |
| FOXO3 | -1.05 | transcription regulator | 2.112 | 2.35E-21 |
| ESR1 | -1.083 | nuclear receptor |  | 7.51E-24 |
| TP53 | 1.263 | transcription regulator |  | 3.26E-43 |
| TP63 | 1.475 | transcription regulator |  | 4.41E-19 |
| IL10 | 1.058 | cytokine |  | 7.55E-21 |
| MYC | 2.057 | transcription regulator |  | 4.67E-21 |
| CDKN1A | 1.69 | kinase |  | 4.11E-24 |
| FAS | 1.442 | transmembrane receptor |  | 1.82E-19 |
| MAPK1 | 1.061 | kinase |  | 3.85E-21 |
| IL10RA | 1.648 | transmembrane receptor |  | 2.66E-21 |

Top 25 based on p-value and shown in order of descending activation z-scores. The Ingenuity Pathway Analysis database lacked enough data to calculate activation z-scores for some canonical pathways. These are left blank.

**Figure 6.**
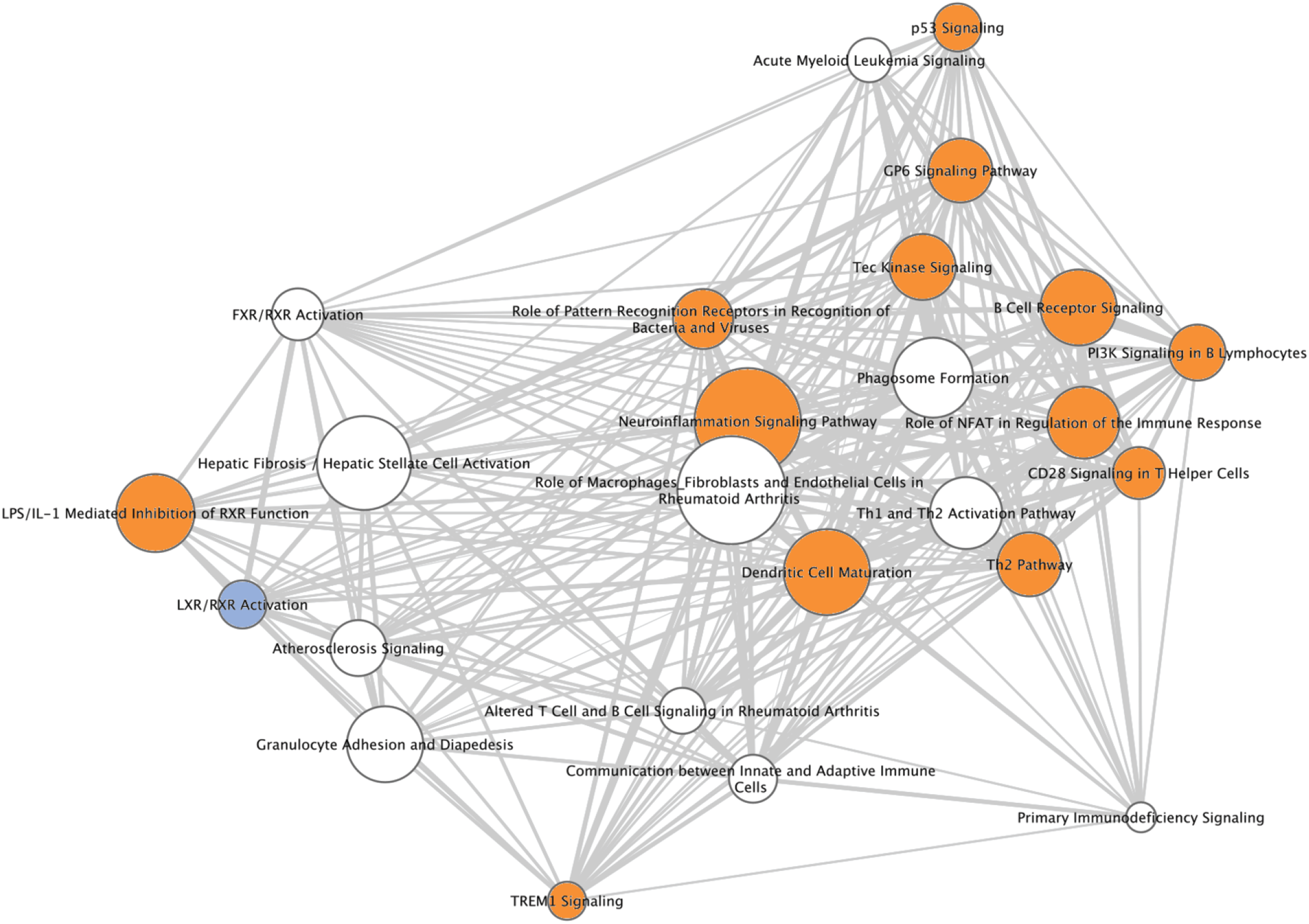
Cluster X pathway network. The top 25 canonical pathways differentially expressed in cluster X were used to generate a network with pathways represented as nodes in the network, with pathways connected by edges. Edges between two pathway nodes indicate that the pathways share at least one gene in common. Nodes are sized according to the number of genes significantly enriched in each pathway and shown as an edge-weighted spring embedded layout with highly related pathways in the center. Nodes with thicker edges between them have higher number of shared genes. Orange and blue colored nodes highlight canonical pathways that are either activated or inhibited, based on an activation Z-score for the pathway, as shown in Table 5. Nodes in white do not have an associated activation Z-score.

A heatmap of cell-specific gene expression values is displayed in Figure 7 with participants grouped by pathology descriptor cluster and genes grouped by cell type. The emerging pattern indicates that the glomerular differential gene expression seen in the cluster comparisons above are occurring in a cell type specific fashion. For example, podocyte enriched genes were downregulated in Cluster X relative to Y+Z whereas monocyte and macrophage enriched genes were upregulated. Furthermore, an influx of monocyte/macrophage cells is seen in cluster X compared to the others, consistent with the observed higher proportion of foam cells (Figure 2B).

**Figure 7:**
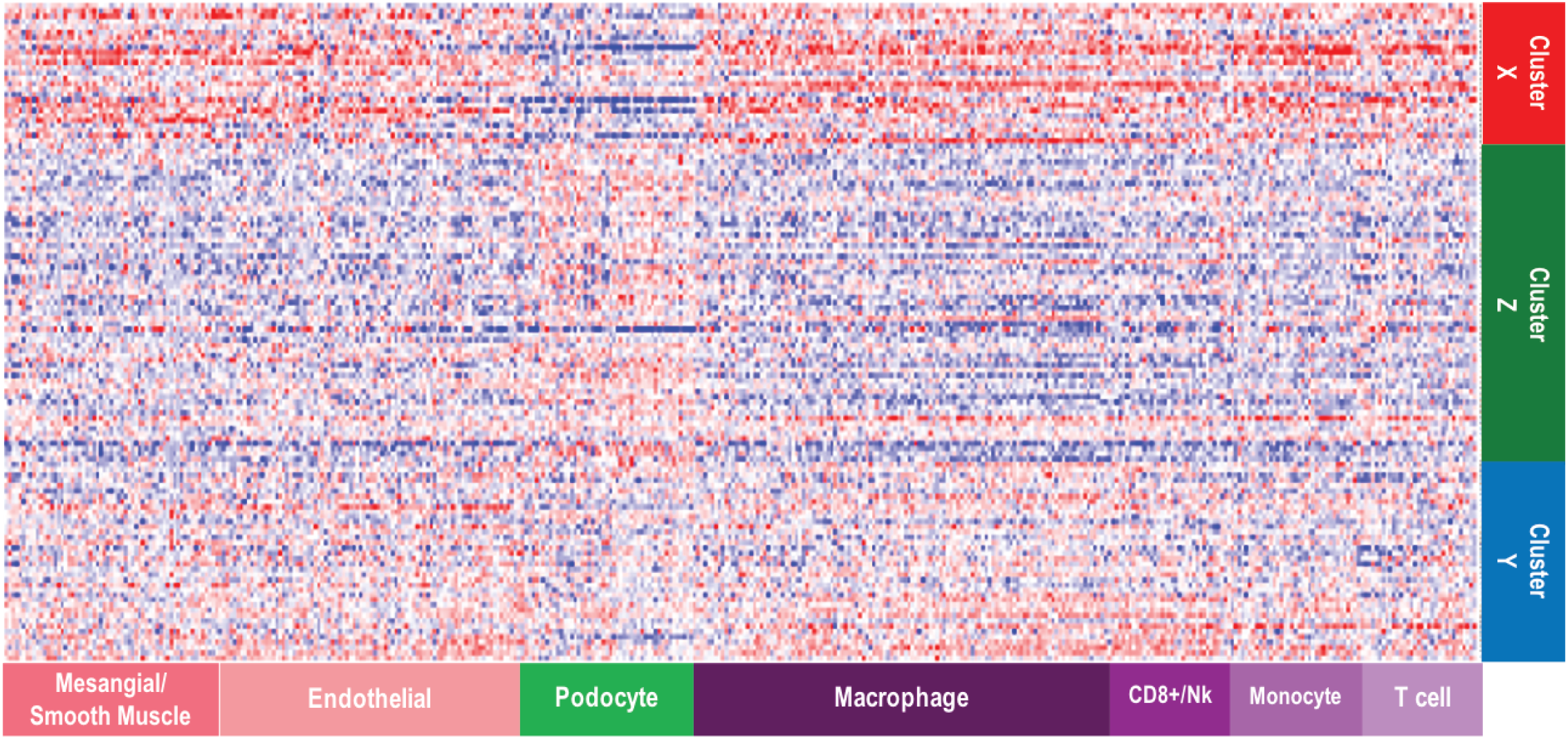
Cell-type specific gene expression across clusters. Cell-type selective gene expression profiles were overlaid onto clustered gene expression data. The expression data were row normalized as Z-scores where blue indicates low expression and red indicates high expression. The emerging pattern indicates that cell-selective gene profiles show evidence of differential gene expression across pathology defined clusters. For example, podocyte-selective gene expression is reduced in Cluster X compared to Clusters Y and Z, whereas macrophage-selective gene expression is increased. This observation strengthens confidence that descriptor clustering is identifying underlying biological mechanism.

Each of the top 15 individual glomerular descriptors shown to drive cluster membership (Table 4) were assessed for correlations with glomerular gene expression. Table 7 shows the canonical pathways and upstream regulators enriched for the seven glomerular descriptors with >700 significant positively and negatively correlated genes. Interestingly, the glomerular descriptor correlated with the most genes was ‘No/Minimal Change’, in the table shown as ‘Any Lesion Seen’, which is simply 100 - percent ‘No/Minimal Change’. Many of the top canonical pathways associated with these gene lists were similar to those associated with Cluster X and enriched for fibrosis, inflammation, and innate immunity pathways. Likewise, top predicted upstream regulators also included cytokines and growth factors TNF, IFNG, IL1B, and TGFB1. The notable exception in Table 7 are the 1401 genes that correlate with the descriptor ‘Segmental Visceral Epithelial Cell Hyperplasia’ (podocytes). Predicted canonical pathways and upstream regulators include those known to be involved with cell growth and regulation of the cell cycle.

**Table 7.**
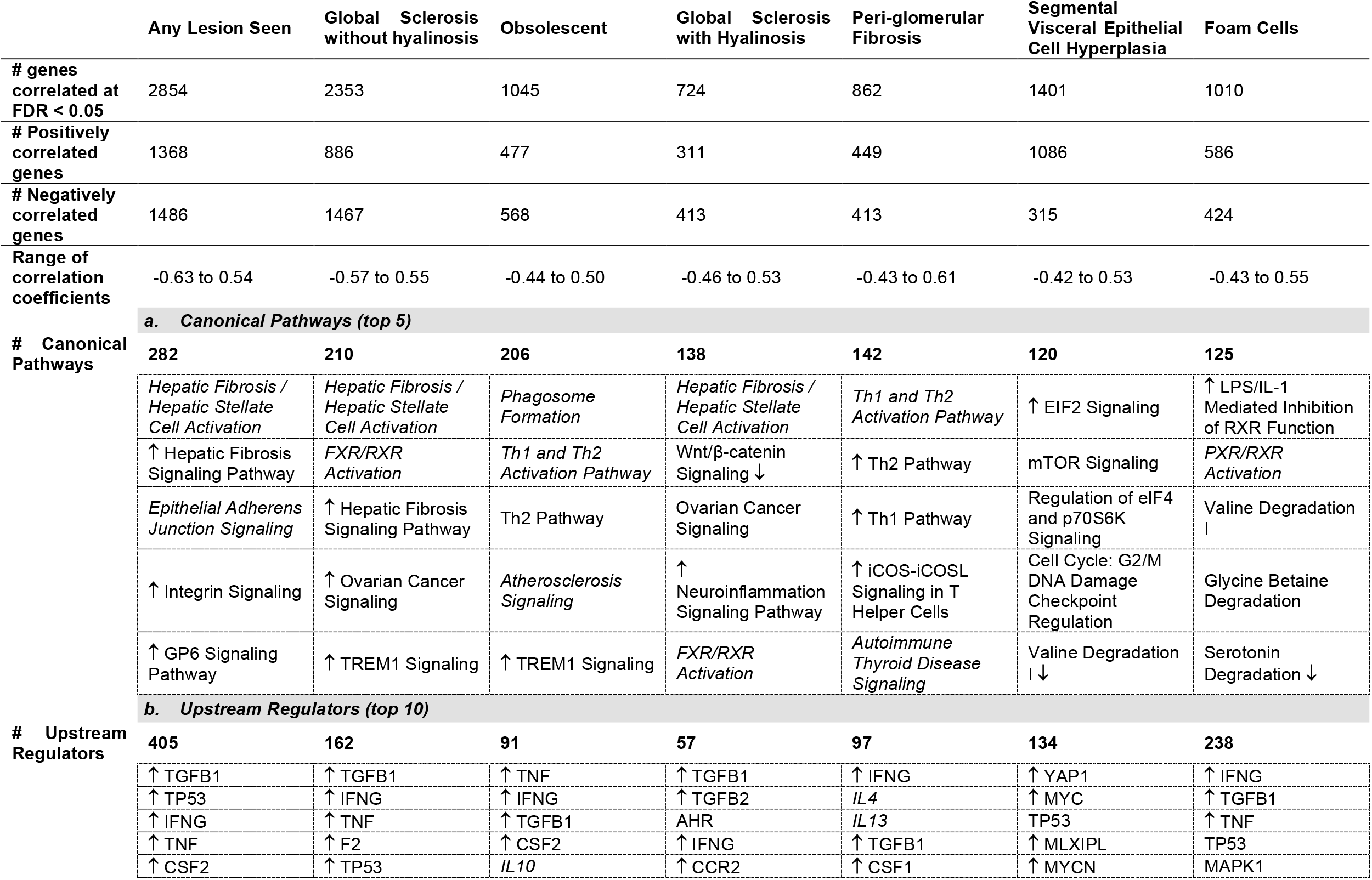
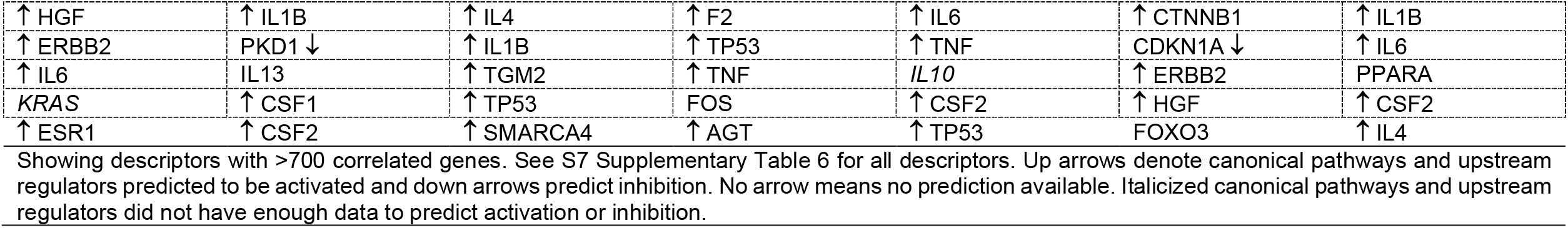
Top (a.) canonical pathways and (b.) upstream regulators predicted from the significant correlation of glomerular RNAseq-derived gene expression with log10-transformed descriptor scores.

|  | Any Lesion Seen | Global Sclerosis without hyalinosis | Obsolescent | Global Sclerosis with Hyalinosis | Peri-glomerular Fibrosis | Segmental Visceral Epithelial Cell Hyperplasia | Foam Cells |
| --- | --- | --- | --- | --- | --- | --- | --- |
| # genes correlated at FDR < 0.05 | 2854 | 2353 | 1045 | 724 | 862 | 1401 | 1010 |
| # Positively correlated genes | 1368 | 886 | 477 | 311 | 449 | 1086 | 586 |
| # Negatively correlated genes | 1486 | 1467 | 568 | 413 | 413 | 315 | 424 |
| Range of correlation coefficients | -0.63 to 0.54 | -0.57 to 0.55 | -0.44 to 0.50 | -0.46 to 0.53 | -0.43 to 0.61 | -0.42 to 0.53 | -0.43 to 0.55 |
| **a. Canonical Pathways (top 5)** |  |  |  |  |  |  |  |
| # Canonical Pathways | 282 | 210 | 206 | 138 | 142 | 120 | 125 |
|  | *Hepatic Fibrosis / Hepatic Stellate Cell Activation* | *Hepatic Fibrosis / Hepatic Stellate Cell Activation* | *Phagosome Formation* | *Hepatic Fibrosis / Hepatic Stellate Cell Activation* | *Th1 and Th2 Activation Pathway* | ↑ EIF2 Signaling | ↑ LPS/IL-1 Mediated Inhibition of RXR Function |
|  | ↑ Hepatic Fibrosis Signaling Pathway | *FXR/RXR Activation* | *Th1 and Th2 Activation Pathway* | Wnt/β-catenin Signaling ↓ | ↑ Th2 Pathway | mTOR Signaling | *PXR/RXR Activation* |
|  | *Epithelial Adherens Junction Signaling* | ↑ Hepatic Fibrosis Signaling Pathway | Th2 Pathway | Ovarian Cancer Signaling | ↑ Th1 Pathway | Regulation of eIF4 and p70S6K Signaling | Valine Degradation I |
|  | ↑ Integrin Signaling | ↑ Ovarian Cancer Signaling | *Atherosclerosis Signaling* | ↑ Neuroinflammation Signaling Pathway | ↑ iCOS-iCOSL Signaling in T Helper Cells | Cell Cycle: G2/M DNA Damage Checkpoint Regulation | Glycine Betaine Degradation |
|  | ↑ GP6 Signaling Pathway | ↑ TREM1 Signaling | ↑ TREM1 Signaling | *FXR/RXR Activation* | *Autoimmune Thyroid Disease Signaling* | Valine Degradation I ↓ | Serotonin Degradation ↓ |
| **b. Upstream Regulators (top 10)** |  |  |  |  |  |  |  |
| # Upstream Regulators | 405 | 162 | 91 | 57 | 97 | 134 | 238 |
|  | ↑ TGFB1 | ↑ TGFB1 | ↑ TNF | ↑ TGFB1 | ↑ IFNG | ↑ YAP1 | ↑ IFNG |
|  | ↑ TP53 | ↑ IFNG | ↑ IFNG | ↑ TGFB2 | *IL4* | ↑ MYC | ↑ TGFB1 |
|  | ↑ IFNG | ↑ TNF | ↑ TGFB1 | AHR | *IL13* | TP53 | ↑ TNF |
|  | ↑ TNF | ↑ F2 | ↑ CSF2 | ↑ IFNG | ↑ TGFB1 | ↑ MLXIPL | TP53 |
|  | ↑ CSF2 | ↑ TP53 | *IL10* | ↑ CCR2 | ↑ CSF1 | ↑ MYCN | MAPK1 |

|  |  |  |  |  |  |  |
| --- | --- | --- | --- | --- | --- | --- |
| ↑ HGF | ↑ IL1B | ↑ IL4 | ↑ F2 | ↑ IL6 | ↑ CTNNB1 | ↑ IL1B |
| ↑ ERBB2 | PKD1 ↓ | ↑ IL1B | ↑ TP53 | ↑ TNF | CDKN1A ↓ | ↑ IL6 |
| ↑ IL6 | IL13 | ↑ TGM2 | ↑ TNF | *IL10* | ↑ ERBB2 | PPARA |
| *KRAS* | ↑ CSF1 | ↑ TP53 | FOS | ↑ CSF2 | ↑ HGF | ↑ CSF2 |
| ↑ ESR1 | ↑ CSF2 | ↑ SMARCA4 | ↑ AGT | ↑ TP53 | FOXO3 | ↑ IL4 |

## DISCUSSION

The introduction of digital pathology provides a platform for the development of new methodologies and protocols for assessment of renal biopsies. The NDPSS capitalizes on digital imaging technology employed in NEPTUNE and facilitates a detailed, quantitative assessment of the renal biopsy.^3^ Reproducibility of the NDPSS has been previously tested ^18^ and optimized;^19^ the majority of descriptors were shown to have good to excellent reproducibility. This granular and quantitative approach, which is agnostic to interpretative diagnoses, offers the opportunity to better explore the relationship between kidney structure, function, and molecular endophenotypes.

The NDPSS is applied to MCD, MCD-like and FSGS patients undergoing their initial, clinically indicated biopsy, and with a broad range of disease severity. The NDPSS is inclusive of a comprehensive list of parameters that can be seen in NS biopsies. Some of these descriptors are employed in current classification schemes (i.e., segmental sclerosis at the vascular pole), but the majority are not (i.e., foam cells, podocyte hypertrophy, periglomerular fibrosis, etc). The clinical and biologic relevance of these parameters, although frequently recognized and occasionally reported by pathologists, was not previously tested, leaving clinicians and pathologists unsure of the prognostic implication of their presence. For example, associations with outcomes were identified for descriptors reflecting global obliteration (such as global sclerosis with and without hyalinosis, global deflation, and obsolescence), periglomerular fibrosis, adhesions, and segmental sclerosis of unknown location. In line with these observations, the value of quantifying global obliteration, independently from the underlying conventional diagnosis, was previously demonstrated in kidney donor biopsies as well as native kidney diseases.^20, 21^ Similarly, descriptors reflecting podocyte hypertrophy and hyperplasia, which were recognized as cellular injury responses in NS,^22, 23^ were identified within the top 15 glomerular descriptors driving cluster membership and were most prevalent in the cluster with worst outcomes (Cluster X). This confirms results from previous morphometric studies in rat models and humans demonstrating that changes in podocyte size and density predict outcome.^24^ A previous study applying the NDPSS to ultrastructural analysis emphasized the importance of a comprehensive analysis to identify previously unrecognized clinically and biologically relevant parameters.^25^ Royal et al.^25^ showed that, although individual podocyte descriptors such as effacement and microvillous transformation were associated with complete remission, endothelial cell abnormalities and groups of patients with the most prominent endothelial cell injury were associated with disease progression and activation of pathways reflecting endothelial cell injury. Thus, the additional information provided by a comprehensive histologic and ultrastructural analysis allows for the identification of patients at highest risk for disease progression and resistance to therapy.

Comprehensively describing kidney histologic changes allows observational data to agnostically define subgroups of patients with morphological similarities (signatures) at the glomerular level and shows that the descriptor-based grouping of patients has clinical relevance. For example, Cluster X contains the highest prevalence of descriptors reflecting global obliteration, segmental glomerulosclerosis of unknown location, periglomerular fibrosis, and podocyte hypertrophy and hyperplasia, while segmental perihilar sclerosis, mid-glomerular sclerosis, and hyalinosis at the vascular pole are most prevalent in Cluster Y. The descriptor-based morphologic signatures and patient clusters were associated with complete remission of proteinuria and loss of kidney function over time, even after adjusting for potential confounding factors and conventional diagnoses. Notably, there was less than full concordance between the descriptor-based clusters and traditional diagnoses of MCD and FSGS variants, highlighting that this systematic and quantifiable approach not only captured the small set of features used as criteria for conventional diagnostics, but also a variety of structural and cellular abnormalities, providing additional information that resulted to be clinically relevant when used to categorize patients with MCD/FSGS. Previous studies focused on the comparison of findings with the conventional disease categories, inferring validity of the results if the findings matched the current classification scheme. This study rather challenges this approach and opens the way for reframing the classification of NS.

Our study links structure and function in FSGS and MCD by revealing associations between descriptor-based clusters and individual descriptors with glomerular gene expression. Previous studies on glomerular specific transcriptomic data from NEPTUNE participants have provided insights into disease molecular mechanisms by highlighting immune activation and Jak-Stat signaling in FSGS,^26^ characterizing expression quantitative trait loci (eQTL),^13^ and integrating with single-cell gene expression to define cellular expression of disease remission in FSGS.^12^ In our analysis we focused on Cluster X to gain insight into the biological and molecular mechanisms associated with the poorest clinical outcomes. A functional analysis of DEGs derived from comparing glomerular gene expression in Cluster X versus Clusters Y+Z revealed significant enrichment for genes in several pathways important for inflammation and immune responses. This is congruent with recent insights into innate immunity activation in the podocyte,^27^ injury to which is key to FSGS and MCD pathogenesis, and current treatment strategies targeting those pathways.^28^ Top predicted causal upstream regulators included TNF, IFNG, IL1B, and TGFB1, which are known to be associated with inflammation and fibrosis.^14-17^

We used single cell RNA sequencing data from reference kidney tissue to highlight cell-type specific gene expression patterns across the three clusters. We found gene expression patterns consistent with decreased steady state expression of podocyte specific genes in Cluster X (the cluster with the worst clinical outcomes) and an increase in monocyte/macrophage specific gene expression. This is consistent with previous observations of podocyte loss or downregulation of podocyte genes with progressive glomerular disease^24^ and the recognition of macrophages and foam cells in FSGS.^29^

By correlating glomerular gene expression with the top glomerular descriptors driving the clusters, we identified individual parameters that may be a surrogate biomarker of mechanism and ultimately a target of novel therapeutic approaches. Seven descriptors correlated significantly with a list of >700 glomerular genes. Genes correlating with ‘Any lesion Seen’ and top descriptors capturing global glomerular obliteration (‘Global Sclerosis with Hyalinosis’, ‘Global Sclerosis without Hyalinosis’, and ‘Obsolescent’) and periglomerular fibrosis demonstrated enrichment for canonical pathways and predicted upstream regulators similar to Cluster X vs Y+Z. Intriguingly, the glomerular descriptor ‘Segmental Viscleral Epithelial Cell Hyperplasia’ correlated with the expression of 1401 glomerular genes enriched for canonical pathways known to be associated with cell hyperplasia, growth, and cell cycle regulation, such as ‘EIF2 Signaling’, ‘mTOR Signaling’, and ‘G2/M Checkpoint Regulation’,^30^ and predicted upstream regulators with key roles in cell growth and gene transcription, including C-Myc, a transcription factor that promotes expression of growth related genes,^31, 32^ and β-catenin, a modulator of cell-cell adhesion and gene transcription.^33^

Our findings support the biological validity of using detailed visual assessment of glomerular pathology because this approach demonstrated correlation with underlying transcriptional programs known to be active in kidney disease. These findings are not definitive; rather they suggest plausible candidate pathways that are potential targets for therapeutic intervention. A final advantage of this approach is the quantifiable data generated by the NDPSS, allowing integration with other omics datasets to understand the biologic relevance of the individual descriptors and patient clusters. These new discoveries can then be studied in animal models or cell cultures to confirm mechanisms and guide development of new treatments for clinical trials.

There are limitations to the approach described for enumerating kidney morphologic changes in glomerular disease tissue. Although we were able to score biopsies from many of participants, the low prevalence of some descriptors may mask their predictive capability using NDPSS. For example, neither glomerular tuft collapse or tip lesions (cellular or sclerosing) were among the top descriptors driving cluster membership, even though they are recognized as features that have poor and good prognoses respectively.^34^ Further, morphologic features seen in cross-section might represent temporal stages of a disease process rather than distinct disease processes. The analysis of follow up biopsies in future studies might yield additional insight into the evolution of glomerular morphology with disease progression. Pediatric patients who do not undergo biopsy were not included in this study; however, this work is paving the way for future work aiming to identify blood or urine biomarkers, which correlate with cluster membership or molecular signatures, and that can be extrapolated to a non-biopsied cohort. Lastly, the current study focused only on histology glomerular descriptors; however, the renal manifestation of glomerular disease processes affects other kidney tissue compartments (tubules, interstitium, and vasculature), which all may contribute to predict outcomes. Previous studies on NEPTUNE datasets, have already shown the predictive ability of scoring for interstitial fibrosis ^35^ and EM descriptors^25^. It is expected that a comprehensive analysis integrating all histologic and ultrastructural descriptors may provide a more complete morphology profile of each biopsy that can better inform clinical outcome prediction.

The present results serve as foundational evidence that comprehensive evaluation of multiple discreet morphologic features from kidney tissue can be used to identify clinically and biologically meaningful glomerular disease categories. Additional work will be necessary to further explore and validate or refine this approach on independent glomerular disease cohorts before implementation in routine clinical practice. Given the rapid evolution of computer-aided image analysis, we anticipate that automated detection, segmentation and quantification of kidney morphologic parameters will be employed toward this goal as an aiding tool to pathologists. Successful development of deep learning models for the segmentation of histologic primitives in the NEPTUNE kidney biopsies has already been demonstrated ^2, 36^, and additional computer-assisted image analysis approaches are being developed. Completion of these future studies could lead to change the current approach for morphologic analysis in clinical practice by implementing interactive human-machine protocols, and data-driven identification of reportable parameters and patient’s categorization, enhancing our ability to better predict outcome and select tailored treatments.

## Data Availability

NEPTUNE gene expression data from microdissected biopsies are available for online interrogation at Nephroseq.org and have been deposited into GEO.

https://www.nephroseq.org/resource/login.html

## SUPPLEMENTARY MATERIAL

**S1.** Supplementary Methods

**S2. Supplemental Table 1**. 37 glomerular descriptors with definitions

**S3. Supplemental Table 2**. Unadjusted associations between each individual glomerular descriptor (n=37) and outcomes

**S4. Supplemental Table 3**. List of 15 differentially expressed genes from comparison cluster X vs Y+Z from RNA sequencing and Affymetrix ST 2.1 platforms

## Acknowledgments

This study was supported by a grant from the National Institute of Diabetes, Digestive, and Kidney Diseases to L.B., J.B.H., B.W.G., L.B.H., L.H.M., and J.Z. (NIDDK R01-DK118431). L.H.M. is supported through funding from NIH/NIDDK, K08 DK115891-01. The Nephrotic Syndrome Rare Disease Clinical Research Network III (NEPTUNE) is part of the Rare Diseases Clinical Research Network (RDCRN), which is funded by the National Institutes of Health (NIH) and led by the National Center for Advancing Translational Sciences (NCATS) through its Office of Rare Diseases Research (ORDR). NEPTUNE is funded under grant number U54DK083912 as a collaboration between NCATS and the National Institute of Diabetes and Digestive and Kidney Diseases (NIDDK). Additional funding and/or programmatic support for this project has also been provided by the University of Michigan, NephCure Kidney International and the Halpin Foundation. All RDCRN consortia are supported by the network’s Data Management and Coordinating Center (DMCC) (U2CTR002818). Funding support for the DMCC is provided by NCATS and the National Institute of Neurological Disorders and Stroke (NINDS).

## NEPTUNE Enrolling Centers

*Cleveland Clinic, Cleveland, OH*: K Dell^*^, J Sedor^**^, M Schachere^#^, J Negrey^#^

*Children’s Hospital, Los Angeles, CA*: K Lemley^*^, E Lim^#^

*Children’s Mercy Hospital, Kansas City, MO*: T Srivastava^*^, A Garrett^#^

*Cohen Children’s Hospital, New Hyde Park, NY:* C Sethna^*^, K Laurent ^#^

*Columbia University, New York, NY:* P Canetta^*^, A Pradhan^#^

*Emory University, Atlanta, GA:* L Greenbaum^*^, C Wang**, C Kang^#^

*Harbor-University of California Los Angeles Medical Center:* S Adler^*^, J LaPage^#^

*John H. Stroger Jr. Hospital of Cook County, Chicago, IL:* A Athavale^*^, M Itteera

*Johns Hopkins Medicine, Baltimore, MD:* M Atkinson^*^, S Boynton^#^

*Mayo Clinic, Rochester, MN:* F Fervenza^*^, M Hogan**, J Lieske^*^, V Chernitskiy^#^

*Montefiore Medical Center, Bronx, NY:* F Kaskel^*^, M Ross^*^, P Flynn^#^

*NIDDK Intramural, Bethesda MD:* J Kopp^*^, J Blake^#^

*New York University Medical Center, New York, NY:* H Trachtman^*^, O Zhdanova**, F Modersitzki^#^, S Vento^#^

*Stanford University, Stanford, CA:* R Lafayette^*^, K Mehta^#^

*Temple University, Philadelphia, PA:* C Gadegbeku^*^, S Quinn-Boyle^#^

*University Health Network Toronto:* M Hladunewich**, H Reich**, P Ling^#^, M Romano^#^

*University of Miami, Miami, FL:* A Fornoni^*^, C Bidot^#^

*University of Michigan, Ann Arbor, MI:* M Kretzler^*^, D Gipson*, A Williams^#^, J LaVigne^#^

*University of North Carolina, Chapel Hill, NC:* V Derebail^*^, K Gibson^*^, E Cole^#^, J Ormond-Foster^#^

*University of Pennsylvania, Philadelphia, PA:* L Holzman^*^, K Meyers**, K Kallem^#^, A Swenson^#^

*University of Texas Southwestern, Dallas, TX:* K Sambandam^*^, Z Wang^#^, M Rogers^#^

*University of Washington, Seattle, WA:* A Jefferson^*^, S Hingorani**, K Tuttle**^§^, M Bray ^#^, M Kelton^#^, A Cooper^#§^

*Wake Forest University Baptist Health, Winston-Salem, NC:* JJ Lin*, Stefanie Baker^#^

*Data Analysis and Coordinating Center*: M Kretzler, L Barisoni, J Bixler, H Desmond, S Eddy, D Fermin, C Gadegbeku, B Gillespie, D Gipson, L Holzman, V Kurtz, M Larkina, J Lavigne, S Li, S Li, CC Lienczewski, J Liu, T Mainieri, L Mariani, M Sampson, J Sedor, A Smith, A Williams, J Zee.

*Digital Pathology Committee*: Carmen Avila-Casado (University Health Network, Toronto), Serena Bagnasco (Johns Hopkins University), Joseph Gaut (Washington University in St Louis), Stephen Hewitt (National Cancer Institute), Jeff Hodgin (University of Michigan), Kevin Lemley (Children’s Hospital of Los Angeles), Laura Mariani (University of Michigan), Matthew Palmer (University of Pennsylvania), Avi Rosenberg (Johns Hopkins University), Virginie Royal (University of Montreal), David Thomas (University of Miami), Jarcy Zee (University of Pennsylvania) Co-Chairs: Laura Barisoni (Duke University) and Cynthia Nast (Cedar Sinai).

*Principal Investigator; **Co-investigator^; #^Study Coordinator

^§^Providence Medical Research Center, Spokane, WA

