## Supplementary methods for "Detailed Quantification of Glomerular Structural Lesions Associates with Clinical Outcomes and Transcriptomic Profiles in Nephrotic Syndrome"

### S1 SUPPLEMENTARY METHODS

#### Study Sample

NEPTUNE is a North American multicenter observational cohort study of children and adults with >0.5 g/day of proteinuria (>1.5 g/day after the first study phase) and a reported diagnosis of MCD, FSGS, or membranous nephropathy. Full NEPTUNE inclusion and exclusion criteria have been previously described.[1] Biopsied participants were enrolled at the time of their clinically-indicated kidney biopsy and a tissue core was obtained for transcriptomic analysis. According to the NDPSS, renal biopsies were processed at the enrolling centers for routine diagnostic work-up, and then collected, de-identified and shipped to the image managing and coordinating center at the National Institutes of Health, where glass slides were scanned to whole slide images (WSI). The WSIs were uploaded and stored, along with biopsy reports, electron microscopy, and immunofluorescence images in a Digital Pathology Repository (DPR).[2] Upon initial review of the WSI by study pathologists for confirmation of the locally rendered diagnosis, MCD was further classified into two subsets: MCD (defined as ≥75% effacement with no global sclerosis or global sclerosis expected for age) or MCD-Like (defined as <75% effacement with or without presence of global sclerosis exceeding that expected for age or ≥75% effacement with global sclerosis exceeding that expected for age).[3] Patients with diagnoses of MCD/MCD-like or FSGS were included in this study.

In accordance with the NEPTUNE study protocol, blood and urine samples were collected during each study visit at 4- to 6-month intervals. Whenever a 24-hour urine sample was not available, a spot urine sample was collected instead. Serum creatinine and 24-hour urine protein (or urine protein to creatinine ratio, UPCR) were measured by a central laboratory. Serum creatinine and UPCR measurements from local laboratories that occurred between study visits were also recorded during the second NEPTUNE study phase. Glomerular filtration rate was estimated using the CKiD-Schwartz equation for children, CKD-Epi for adults over 26 years old, and an average of the two equations for adults between 18-26 years old.[4] Clinical data, including demographics, medical history, kidney failure status, medication exposures and local laboratory results, were collected at each study visit.

**NEPTUNE Digital Pathology Scoring System (NDPSS)**

A standardized protocol for annotation (enumeration) of glomeruli through all biopsy levels was employed.[2, 5] Annotated glomeruli were then scored by study pathologists using the NDPSS. A descriptor reference manual to guide the scoring process and an electronic scoring sheet was used to record the scoring of 37 glomerular descriptors through all biopsy sections and stains (Figure 1) (Table S1: list of descriptors and definitions).[2, 6] Previous studies have already demonstrated good to excellent reproducibility with the majority of NDPSS descriptors.[7, 8]. The overall presence of each descriptor in a participant's biopsy was quantified by calculating the percentage of a participant's glomeruli with each descriptor present.

**Patient Clustering by Morphologic Profiles**

Each descriptor was first scaled to its observed range—for example, a descriptor that was only observed in 0-15% of glomeruli across all participant's biopsies and another descriptor that was observed in 0-100% of glomeruli were both scaled to 0-1. We also conducted a sensitivity analysis using unscaled data. Using the 37 glomerular descriptors, we applied Ward's hierarchical clustering algorithm to cluster participants into subgroups representing similar biopsy profiles. Hierarchical clustering produces a dendrogram, or tree diagram, showing groupings (branches) of similar participants and how different numbers of clusters can be created depending on how branches are trimmed. To determine the optimal number of clinically relevant clusters, we started with two clusters and assessed whether the two clusters were different in terms of clinical outcomes based on an unadjusted Cox proportional hazards model. If the two clusters were associated with different clinical outcomes, we determined whether three clusters were appropriate by assessing whether the one cluster that split into two had different clinical outcomes. We then repeated this process until we did not find evidence of a difference in clinical outcomes. The final cluster solution was compared with conventional morphologic classifications (MCD/MCD-Like and FSGS subtypes) [9, 10] using a Sankey diagram.

To identify the strongest individual descriptors driving cluster membership, we used elastic net penalized multinomial regression with a penalization mixing parameter of $\alpha=0.01$. Prediction strength of each descriptor was determined based on order of entry into the model by varying the shrinkage parameter $\lambda$ from very large (few descriptors would enter into the model) to very small (all descriptors would be in the model). We also assessed whether a subset of descriptors could predict cluster membership by repeating the model with only the top 5, 10, 15, 20, 25 and 30 descriptors and comparing prediction accuracy across models.

**Clinical Outcomes Analysis**

Clinical outcomes of interest included **(1)** time from biopsy to complete remission of proteinuria, defined as UPCR <0.3 mg/mg, **(2)** time from biopsy to a composite disease progression outcome of at least 40% decline in eGFR [11] with eGFR <90 mL/min/1.73$m^2$ or kidney failure, and **(3)** longitudinal eGFR over time. For the two time-to-event outcomes, we used Cox proportional hazards models, and for longitudinal eGFR, we used a linear mixed model with random intercept and random slope for each participant to account for repeated measures. Interactions between time from biopsy and cluster groups were tested in the longitudinal eGFR model to assess whether eGFR rate of change (i.e., slope) differed across clusters.

We performed both unadjusted and sequentially adjusted models between cluster groups and outcomes to evaluate whether associations between cluster groups and outcomes would be affected by covariate adjustments. For adjusted models, we first adjusted for demographics and clinical characteristics. If outcomes had sufficient numbers of events or observations to avoid overfitting, we adjusted for age, sex, self-reported black race, Hispanic ethnicity, eGFR at biopsy, UPCR at biopsy, kidney disease duration as of biopsy, and any immunosuppression use at or within 30 days prior to biopsy; if there were not enough events or observations to avoid overfitting, we used backward selection to choose adjustment covariates. Then, we additionally adjusted for participant disease diagnosis (MCD/MCD-Like vs. FSGS). Finally, we used unadjusted models to assess associations between each individual glomerular descriptor and each outcome. P-values were controlled for false discovery rate (FDR) using the Benjamini and Hochberg linear step-up method.

**Molecular Data and Analysis**

To determine whether the descriptor-based clusters were associated with specific biological processes, transcriptomic analysis was performed on kidney biopsy samples. Microdissected glomerular RNA sequencing (RNA-seq) expression data were available for a subset of NEPTUNE participants with biopsy-proven MCD/MCD-Like or FSGS. Briefly, mRNA samples were prepared using the Illumina TruSeq mRNA Sample Prep v2 kit. Multiplex amplification was used to prepare cDNA with a paired-end read length of 100 bases using an Illumina HiSeq2000 at the University of Michigan Advanced Genomics Core (brcf.medicine.umich.edu/cores/advanced-genomics/). Quality of the sequencing data was assessed using the FastQC tool (www.bioinformatics.babraham.ac.uk) and read counts were extracted from the fastq files using HTSeq (version 0.11). Each of the identified clusters was compared to the remaining clusters using Significance Analysis of Microarray (SAM). Differentially expressed genes (DEGs) were defined as having fold change of greater than 1.3 and false discovery rate (FDR)-corrected q-value less than 5%. DEGs were analyzed for enrichment of canonical pathways and upstream regulators using the Ingenuity Pathway Analysis Software Suite.[12] An activation z-score was derived from the direction of gene regulation observed to the expected direction of gene regulation to infer the activation state of a putative upstream regulator,[12] with adjustment for multiple testing using Bonferroni less than 5% as the criterion for statistical significance. Upstream regulators were identified using both enrichment as well as direction of expression change with known cause and effect relationships.

To assess whether biological processes were associated with the individual descriptors driving cluster membership, a Pearson correlation coefficient was calculated between each glomerular RNA-seq-derived gene expression value and log10-transformed descriptor percentage score (after descriptors with 0% were transformed to 1% to avoid undefined values), with adjustment for multiple testing using q-value less than 5% as the criterion for statistical significance. Significantly correlated transcripts were analyzed for enrichment of canonical pathways and upstream regulators using the Ingenuity Pathway Analysis Software Suite.[12] An activation z-score was derived from the direction of correlation relative to expected direction of gene regulation to infer the activation state of a putative upstream regulator,[12] with adjustment for multiple testing using Bonferroni less than 5% as the criterion for statistical significance.

To understand the heterogeneity of kidney cell types in each pathology cluster, we generated a list of genes that are specific to known renal and non-renal cell types using single cell RNA-seq profiles derived from three tumor nephrectomy datasets, a subset of data from Menon et al.[13] The expression values of the subset of cell-specific genes were extracted from the glomerular compartment bulk transcriptome data and were labeled according to cell type.[14] A heatmap was generated based on the VOOM transformed log expression in which participants were ordered based on the pathology cluster assignment and transcripts were grouped by cell type. The heat map generated visualizes the cell type specific transcript expression pattern across the three pathology clusters.

Of the 126 descriptor scored cases that had glomerular RNA sequencing data available, 77 also had transcriptomic profiling by the Affymetrix ST 2.1 platform. Glomerular gene expression from cluster X (N=14) was compared to the clusters Y+Z (N=63) using Significance Analysis of Microarray (SAM). Differentially expressed genes (DEGs) were defined as having fold change of greater than 1.3 and false discovery rate (FDR)-corrected q-value less than 5%.
